# Effective Deep Learning Approaches for Predicting COVID-19 Outcomes from Chest Computed Tomography Volumes

**DOI:** 10.1101/2020.10.15.20213462

**Authors:** Anusua Trivedi, Anthony Ortiz, Jocelyn Desbiens, Caleb Robinson, Marian Blazes, Sunil Gupta, Rahul Dodhia, Pavan Bhatraju, W. Conrad Liles, Aaron Lee, Juan M. Lavista Ferres

## Abstract

The rapid evolution of the novel coronavirus SARS-CoV-2 pandemic has resulted in an urgent need for effective clinical tools to reduce transmission and manage severe illness. Numerous teams are quickly developing artificial intelligence approaches to these problems, including using deep learning to predict COVID-19 diagnosis and prognosis from computed tomography (CT) imaging data. In this work, we assess the value of aggregated chest CT data for COVID-19 prognosis compared to clinical metadata alone. We develop a novel patient-level algorithm to aggregate the chest CT volume into a 2D representation that can be easily integrated with clinical metadata to distinguish Novel Coronavirus Pneumonia (COVID-19+) from other cases of viral pneumonia and normal healthy chest CT volumes with state-of-the-art performance. Furthermore, we present a multitask model for joint segmentation of different classes of pulmonary lesions present in COVID-19 infected lungs that can outperform individual segmentation models for each task. We directly compare this multitask segmentation approach to combining feature-agnostic volumetric CT classification feature maps with clinical metadata for predicting mortality. These approaches enable the automated extraction of clinically relevant features from chest CT volumes for risk stratification of COVID-19+ patients.

## 1 Introduction

The Coronavirus Disease (COVID)-19 pandemic has generated an unprecedented global health response in an effort to reduce transmission and mortality. Since the early stages of the pandemic, computed tomography (CT) chest imaging has been a valuable assessment tool. Experts have developed an understanding of COVID-19-associated chest CT findings, which include ground-glass opacities (GGOs), consolidation, bilateral involvement, and peripheral and diffuse distribution (Lee, Ng, and Khong 2020). Early chest CT abnormalities may be absent or nonspecific; however (Bernheim et al. 2020; Shi et al. 2020b) and deep learning models have been developed to identify subtle imaging features and distinguish COVID-19 pneumonia from normal findings or other pneumonias.(Li et al. 2020) Deep learning models have also been trained to predict outcomes such as hospitalization, intubation, and/or mortality based on CT imaging data.(Wang et al. 2020; Xiao et al. 2020)

Despite these advances, machine learning approaches with volumetric CT data remain challenging due to a large number of slices in relatively few patients (the curse of dimensionality). Many advances rely on using each slice as independent training examples. In this study, we explore deep learning approaches for extracting clinically relevant features from chest CT volumes using limited data. We develop a novel method for aggregating the chest CT volume into a 2D representation of global features that can distinguish Novel Coronavirus Pneumonia (COVID-19+) from other viral pneumonias and normal chest CT volumes, with state-of-the-art performance. Furthermore, we present a multitask model for joint segmentation of different classes of pulmonary lesions present in COVID-19 infected lungs with the goal of extracting local, highly relevant features. We then create prognostic models using the extracted features together with patient demographic data, comparing the performance of models using different combinations of relevant data to predict mortality. The overall goal of this work is to enable automated extraction of relevant features from chest CT volumes that can be incorporated with clinical data for risk stratification of COVID-19+ patients. Our contributions include:

- Presenting two novel approaches for extracting clinically relevant features from limited chest CT imaging data.
- Demonstrating that resulting features improve the performance of prognostic models when combined with other clinical data.

## 2 Related Work

### Classification

There are already some published studies on CT based COVID-19 diagnosis systems (Shi et al. 2020a; Dong et al. 2020). (Zhang et al. 2020; Al-karawi et al. 2020) created COVID-19 analysis framework on a dataset comprising 4,154 patients, and it can separate COVID-19 from other basic pneumonia. (Li et al. 2020) built an AI framework for COVID-19 identification on a dataset comprising of 3,322 subjects. (Zhang et al. 2020; Al-karawi et al. 2020; Li et al. 2020; Xiao et al. 2020; Shi et al. 2020b) proposed different feature extraction approaches to exploit the power of CT scans for COVID-19 diagnosis and prognosis. (Li et al. 2020) developed a 3D deep learning model for the diagnosis of COVID-19, referred to as COVNet. COVNet takes as input a series of CT slices, extract features from each slice using a ResNet50 backbone, and finally combine obtained features using the max-pooling operation. Obtained features are then used to generate a classification prediction for the entire CT scan. (Zhang et al. 2020) process CT scans in a two steps fashion.

- **2D**: 2D quantitative properties including lung area as the total count of pixels not belonging to the background, lesion area as the total count of pixels presenting lung lesions, where lesions are obtained for each CT slice through semantic segmentation models.
- **½D**: Volume features are obtained through the integration of all 2D-size based quantitative properties of CT slices.

(Al-karawi et al. 2020) use Fourier processing of the scan slices to obtain its FFT-spectrum from which a texture type is deduced and an appropriate feature vector is extracted for further ML analysis.

### Segmentation

In addition to diagnostic models, several prediction models have been proposed based on an assessment of lung lesions. There are three typical classes of lesions that can be detected in COVID-19 chest CT scans: ground-glass opacity (GGO), consolidation, and pleural effusion (Shi et al. 2020c; Ng et al. 2020). Imaging features of the lesions, including shape, location, extent, and distribution of involvement of each abnormality, have been found to have good predictive power for mortality (Yuan et al. 2020) or hospital stay (Qi et al. 2020). These features, however, are mostly derived from the delineated lesions, and so depend heavily on lesion segmentation. Harrison et al. (Harrison et al. 2017) showed that a deep learning-based segmentation beats a specialized methodology in cases with interstitial lung maladies. Following the same idea, automatic lung lesion segmentation for COVID-19 has been actively investigated in recent studies. (Shan et al. 2020) proposed a VB-Net-based neural network to segment the infection regions in CT scans. This model, when trained using CT scans of 249 COVID-19 patients, achieved a Dice score of 0.92 between automatic and manual segmentation, and successfully reduced the delineation time to but 4 minutes. In another recent study by (Chaganti et al. 2020), a lesion segmentation model supported the 3D-Dense U-Net architecture was proposed and trained on CT scans of a mixture of 160 COVID-19, 172 viral infection, and 296 interstitial lung disease patients. Although the lesion masks weren’t compared voxel-to-voxel, the volumetric measures of lesions, like percentage of opacity and consolidation, showed a high correlation between automatic and manual segmentation. These studies suggest that lesion features could be a useful biomarker for COVID-19 patient severity assessment.

### Prognosis

Prognostic models based on automated lesion segmentation features have also been developed. (Xiao et al. 2020) created a deep learning model using multiple instance learning and ResNet34 to analyze CT images from 408 COVID-19+ patients for severe disease (AUC of 0.892, CI: 0.828–0.955). The model was able to predict severe disease in a subgroup analysis of patients who presented with non-severe symptoms. Other studies have combined CT features with other clinical data to predict outcomes. (Zhang et al. 2020) uses volumetric lung lesion features extracted from the segmentation model along with clinical metadata. They combine the features using a tree-based ensemble model to predict disease severity (defined as ICU admit, intubation, or death), and show that the pulmonary lesions were most predictive for progression to severe disease, followed by clinical parameters related to lung function, age, and fever on admission. (Wang et al. 2020) used CT images to train a model to diagnose COVID-19 and risk-stratify patients for severe disease. The first model segmented the lung region then performed a non-lung area suppression operation to hide any non-pulmonary features further. The prognostic model was first trained on a large dataset of 4106 CT-EGFR+ lung cancer patients to predict EGFR mutation status (wild type vs. mutant) based on the segmented lung region and subsequently trained to detect COVID-19 in a separate dataset of 1266 patients (of which 924 were COVID-19+). A set of three COVID-19 features were extracted and combined with clinical data (age, sex, comorbidities) to classify patients into high-risk/low-risk categories. The patients classified as high risk by the model were shown to have significantly longer hospital stays.

## 3 Intensity Map Projection for Classification

We first present a solution for global feature extraction that falls into the 2½D model category and relies heavily on the so-called texture analysis theory.

### 3.1 Texture Analysis

Texture analysis theories and methods have proven helpful in describing medical images. The oldest and most widely used texture analysis technique is the intensity histogram. It counts the number of occurrences of intensity values for all pixels of an image and builds a histogram from the cumulative entries. Its output is also a set of features/frequencies.

#### Occlusion Mask

Sometimes, partly visible objects on an image are blocked by other objects located in the foreground. This phenomenon is called *occlusion*. One can think of lesions (GGO, consolidations, fibrosis, …) as some kind of occlusion objects masking the “normal” background image.

Let *f*_*k*_, with *k* = 0, …, *N* − 1, be *N* images, or planes, of the same size. Then, the occlusion mask set of the *f*_*k*_ planes with respect to a given *label* function *ϕ* : *χ* → ℤ is 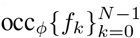. In other words, for any pixel *x*, the label *ϕ*(*x*) determines which one of the multiple pixel values 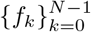 actually appears in the final image 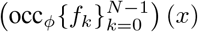 at precisely *x*.

#### Composite Image

Let 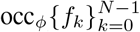 be an occlusion mask set with *N* planes. Then, the expected value of the histogram transform of a composite image (the original image) is a linear convex combination of the histograms of each individual plane, *i*.*e*.

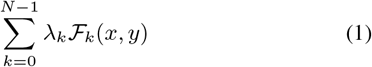

where 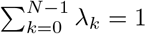, and *λ*_*k*_ ≥ 0 for all *k* = 0, …, *N* − 1. Here, the ℱ_*k*_ are the histograms of the *f*_*k*_ planes, for *k* = 0, …, *N* − 1 (See (Massar et al. 2013), Theorem 1).

#### Volume Decomposition

Let *v* a 3D-volume composed of *M* horizontal planes *p*_*j*_, and let *π*_*j*_(*x, y, z*) = (*x, y*) be the projection from *v* over *p*_*j*_, for all *j*. The *p*_*j*_ are linear transformations. Put *v* = *p*_0_ ⊕ *p*_1_ ⊕ … ⊕ *p*_*M*−1_, with ⊕ being the image stacking operation (see Fig. 2b). If 𝒱 is the histogram of *v*, and 𝒫_*j*_ is the histogram of *p*_*j*_ = *π*_*j*_, for *j* = 0, …, *M* − 1, then we have, by virtue of the linearity of the projections *π*_*j*_,

**Figure 1:**
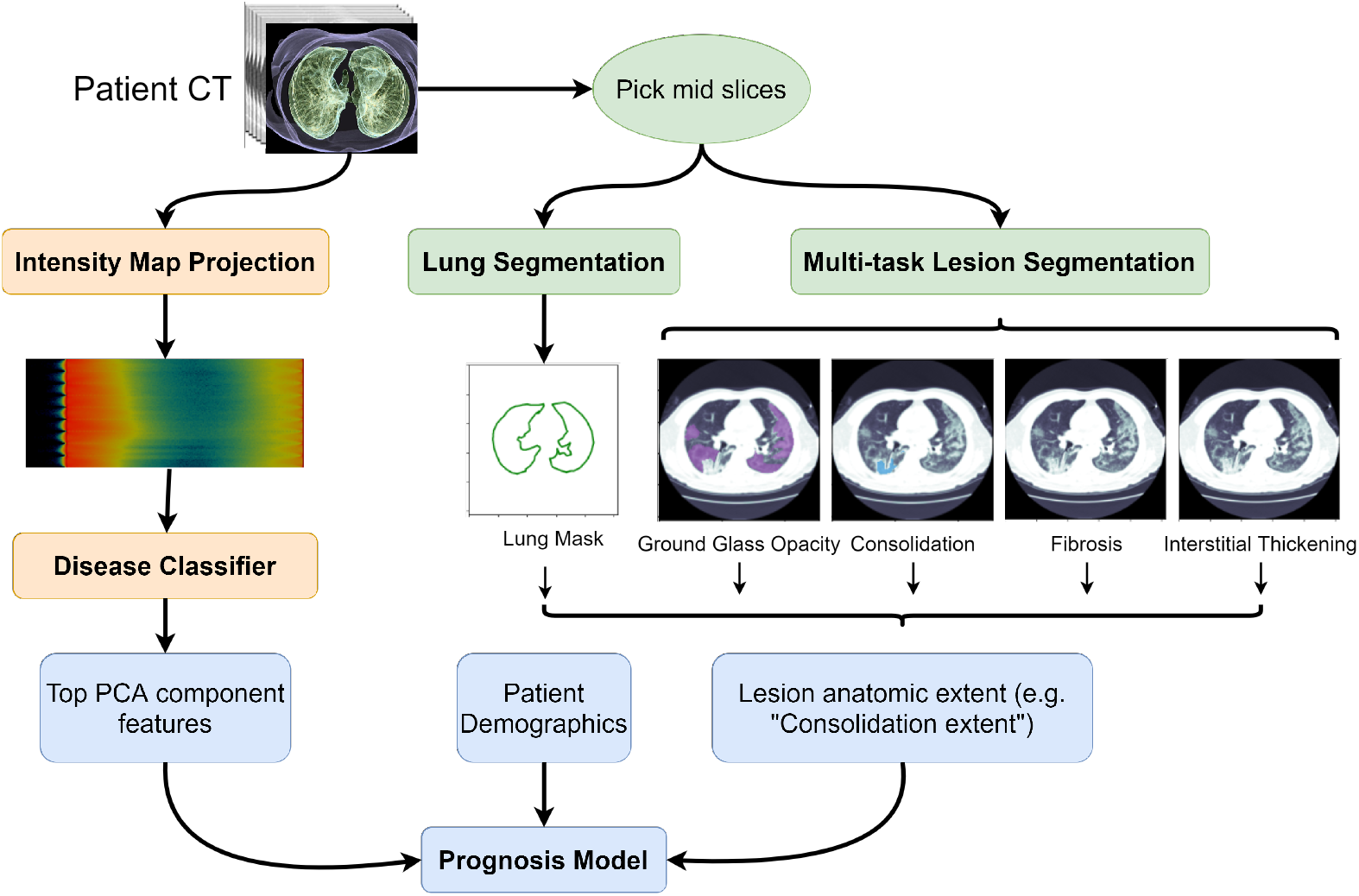
CT-based machine learning pipeline for COVID-19 prognosis. The left side of the figure (orange) represents the the intensity map projection and disease classifier presented in Section 3. The right side if the figure (green) show the use of multitask semantic segmentation to obtain lesion anatomic extent features. Features obtained from the disease classifier, lesion anatomic extent features, and patient’s demographics are then used for mortality prediction using a prognosis model (blue).

**Figure 2:**
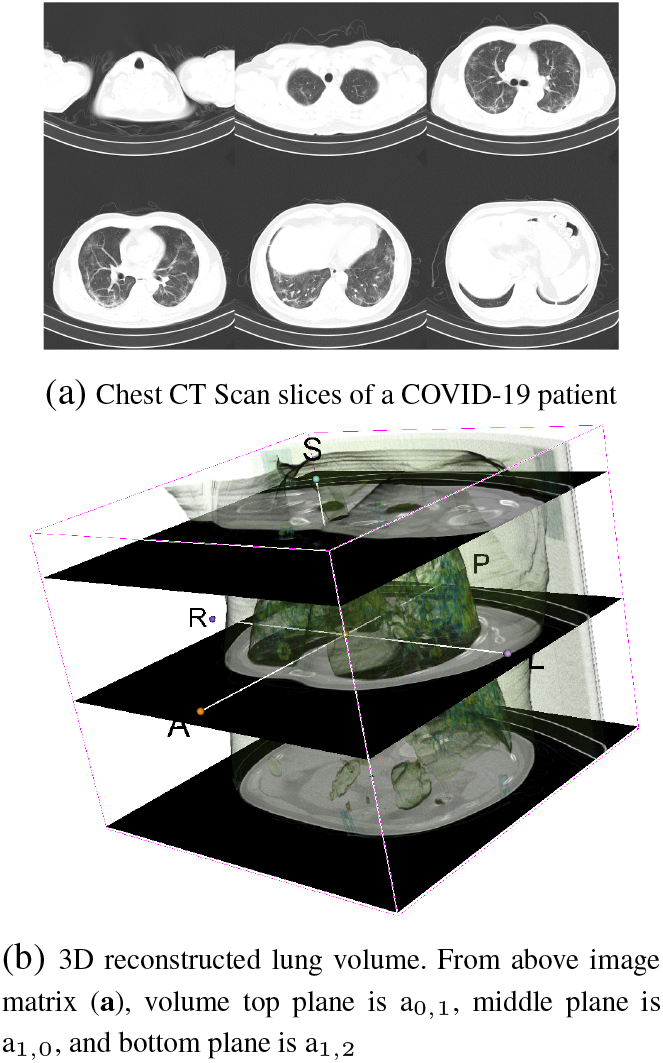
CT Scan slices and lung volume reconstruction

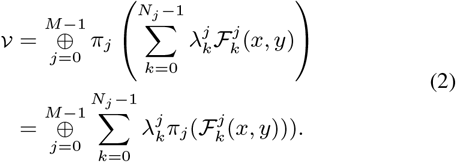

In conclusion, and under the operator ⊕, the histogram of a volume *V* is the sum of the histograms of its projections over the composing planes *p*_*j*_, with the convention that lesions are mapped to the occlusion mask sets 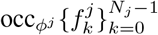 for *j* = 0, …, *M* − 1.

### 3.2 Intensity Map

Based on Eq. 1 and Eq. 2, we propose to aggregate individual CT slices/planes for creating the *intensity map* as shown in Algorithm 1.

#### Algorithm 1 Intensity Map Projection

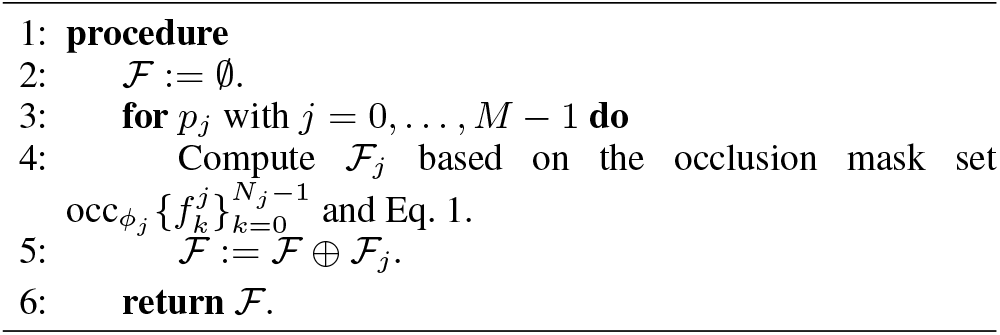

The algorithm sets the CT scan as the unit of prediction while preserving the natural ordering of the slices/planes in the final 2D intensity map.

As an example, Fig. 3 is the calculated intensity map of the data shown in Fig. 2. Red color means low intensity, while blue color means high intensity. COVID-19 maps are bluer than Pneumonia/Normal maps because they host on average more high-intensity pixels than the average normal plane.

**Figure 3:**
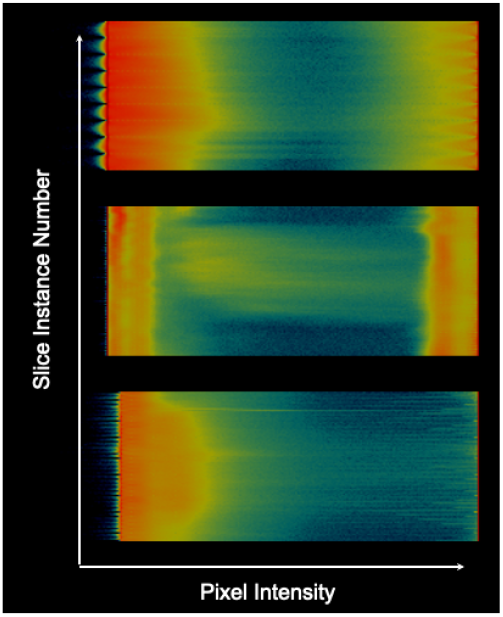
Intensity maps. Top: Normal, Middle: Pneumonia, Bottom: COVID-19+. Red color means low intensity, blue color means high intensity, and black no intensity.

### 3.3 Diagnosis Experiments on CC-CCII Dataset

#### CC-CCII Dataset

CC-CCII dataset is a large CT dataset from the China Consortium of Chest CT Image Investigation (CC-CCII), consisting of CT images from retrospective cohorts. It consists of CT images from Sun Yat-sen Memorial Hospital and Third Affiliated Hospital of Sun Yatsen University, The first Affiliated Hospital of Anhui Medical University, West China Hospital, Guangzhou Medical University First Affiliated Hospital, Nanjing Renmin Hospital, Yichang Central People’s Hospital, Renmin Hospital of Wuhan University. CT imaging was performed as a part of patients’ routine clinical care, including CT images from novel coronavirus pneumonia (COVID-19+) and other viral common pneumonia (Pneumonia). Pneumonia group consists of common types of viral pneumonia, including adenoviral, influenza, and para-influenza pneumonia.

The classification dataset we used in this paper is a cleaned subset of the CC-CCII (Zhang et al. 2020) dataset. It consists of a total of 514,103 CT slices from 2,471 patients. It includes 156,070 slices from 839 COVID-19+ patients, and 159,700 slices from 874 viral Pneumonia patients. Normal dataset counts for 95,459 slices for a total of 758 patients.

#### Dataset Breakdown

We split the cleaned out version of the CC-CCII dataset previously described as explained in Table 1.

**Table 1:** Per-Patient Train/Validation/Test dataset splits

| # of Patients | Train | Validation | Test |
| --- | --- | --- | --- |
| Pneumonia | 774 | 50 | 50 |
| COVID-19+ | 739 | 50 | 50 |
| Normal | 658 | 50 | 50 |

For each CT scan, we created an intensity map representation following the methodology described in the previous section. The intensity maps can then be used as input to a classification model.

#### Classification Model

We used InceptionResnetV2 model trained on 256 *×* 256 intensity map images. We used Nadam optimizer with a batch size of 12. Learning rate was set to 0.0001. Trained was done over 96 epochs. Categorical Cross-Entropy was used as the loss function. The classification model was developed using Keras.

### 3.4 Diagnosis Results on CC-CCII Dataset

Table 2 shows the achieved performance in both validation and test sets on the CC-CCII dataset.

**Table 2:** Per-Patient Validation/Test results

| Performance | Pneumonia | COVID-19+ | Normal |
| --- | --- | --- | --- |
| Accuracy (%) | 98.0/98.0 | 95.3/97.3 | 97.3/96.7 |
| Area under ROC | 97.0/98.0 | 96.0/98.5 | 96.5/95.5 |
| Specificity (%) | 100.0/98.0 | 94.0/94.0 | 99.0/99.0 |
| Sensitivity (%) | 100.0/98.0 | 94.0/97.0 | 99.0/99.0 |
| F1 Score (%) | 96.9/97.0 | 93.3/96.1 | 95.9/94.8 |

On a per-patient basis, the overall accuracy of the InceptionResnetV2 classifier trained using our intensity map projection is 95.3% or the validation set and 96.0% on the test set with 95% confidence interval of (91.958, 98.709)% and (92.864, 99.136)% respectively.

On a per-scan basis, our overall accuracy is 97.3% on the test set and 97.10% on the validation set with 95% CI of (95.911, 98.634)% and (95.686, 98.495)% respectively.

Table 4 from (Zhang et al. 2020) is the reference baseline generated over the same dataset on a per-scan basis. In this case, overall accuracies were 92.49%/89.92% while COVID-19+ accuracy = 92.49%/90.70%, sensitivity = 94.9%/92.51%, specificity = 91.13%/85.92%, and ROC = .9797/.9712.

**Table 3:** Per-Scan Validation/Test results

| Performance | Pneumonia | COVID-19+ | Normal |
| --- | --- | --- | --- |
| Accuracy (%) | 97.6/98.2 | 98.7/98.0 | 98.2/98.0 |
| Area under ROC | 99.0/100.0 | 100.0/100.0 | 100.0/100.0 |
| Specificity (%) | 98.0/98.0 | 99.1/98.6 | 98.7/99.0 |
| Sensitivity (%) | 97.0/98.5 | 98.0/97.0 | 96.7/95.3 |
| Score (%) | 96.8/97.5 | 98.2/97.2 | 96.7/96.3 |

**Table 4:** Internal/External validation results (Zhang et al. 2020)

| Performance | Pneumonia | COVID-19+ | Normal |
| --- | --- | --- | --- |
| Area under ROC | 96.7/96.8 | 98.0/97.1 | 99.5/99.9 |

Comparing Table 3 with Table 4, we may say that our method achieves higher AUC value than the baseline method for the three classes Pneumonia/COVID-19+/Normal. COVID-19+ sensitivity/specificity/ROC scores are also better showing the usefulness of our proposed representation. Moreover, our method does not rely on lung or lesion segmentation, making *a fortiori* the economy of one classifier. Very few other studies use full scans for their research due to the lack of publicly available data. Most existing COVID-19 ML models using CT scans are built upon a handful of selected slices displaying visible lesions.

Our final objective is to accurately perform prognosis on COVID-19 positive patients using CT scans. As shown above, our intensity map projection provides a great representation to learn diagnosis models. However, it removes local spatial information, such as pulmonary lesions, which has been proven to be important for prognosis. In the following section, we propose a multitask segmentation network to extract additional features needed for prognosis.

## 4 Obtaining Pulmonary Lesion features for COVID-19 Prognosis

After a patient is confirmed as being COVID-19 positive using the classification network, we can use segmentation strategies to quantify pulmonary lesions in patients’ lungs. Accurate segmentation of these lesions and their percentage of lung coverage can facilitate the estimation of the severity of the disease and help improve prognosis on patients. We propose a pipeline to accurately compute the lesion’s lung coverage on COVID-19 patients using a multitask lesion segmentation network along with a lung segmentation network.

### 4.1 Problem Abstraction

We let 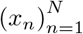 represent a set of training CT slice images. Each slice *x*_*n*_ is associated with a corresponding lung contour mask *l*_*n*_. Depending on whether the slice *x*_*n*_ shows the presence of a pulmonary lesion it will be also associated to the corresponding lesion masks element of the set *lesions* ∈ {*ggo*_*n*_, *cl*_*n*_, *fl*_*n*_, *it*_*n*_ }. For each pixel (*i, j*) in CT slice *x*_*n*_ we aim to assign a label *l*_*n*_ when the pixel is located within the lung contours, *ggo*_*n*_ for pixels where ground glass opacity is present, *cl*_*n*_ for consolidation, *fl*_*n*_ for every pixel showing signs if pulmonary fibrosis, and *it*_*n*_ for pixels where interstitial thickening is present. The lesion coverage for a particular lesion can then be computed as the ratio between the total amount of pixels showing signs of the lesion and the total of lung contour (*l*_*n*_) pixels present on the slice. It is important to notice that a pixel (*i, j*) in a CT slice image can be associated with multiple lesions.

#### CC-CCII Pulmonary Lesions Segmentation Dataset

A subset of 1290 cleaned up CT slices were obtained from a set of manually segmented slices from the CC-CCII dataset. The annotation was done via polygons. The segmentation labels were selected as relevant pathological features for distinguishing COVID-19+ and other common pneumonia. The annotation included lung field, and five commonly seen categories of lesions, including lung consolidation (CL), ground-glass opacity (GGO), pulmonary fibrosis, interstitial thickening and pleural effusion. Segmentation results were annotated and reviewed by five senior radiologists with 15 to 25 years of experience.

#### Experimental Setup and Evaluation Metrics

The available segmentation slices were randomly assigned to three distinct sets: training (80%), validation (10%), and test (10%). Models are evaluated on the held-out test set.

We report performance results using mean intersection over union (mIoU), a standard metric for semantic segmentation. (Long, Shelhamer, and Darrell 2015)

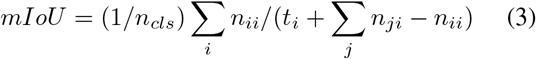

where *n*_*ij*_ is the number of pixels of class i predicted to belong to class j, there are *n*_*cls*_ different classes, and *t*_*i*_ = Σ_*j*_ *n*_*ji*_ is the total number of pixels of class i.

Some lesion classes are not present in the slice. In those cases we assume that background is the only class and assign mIoU of 1 to models not predicting that class. This make this metric not very informative for certain lesions. Hence, we also report Global mean IoU (GIoU) as the mean IoU calculated only over slices where the lesion of interest is present.

### 4.2 Multi-task Network for Segmentation of Pulmonary Lesions in COVID-19 patients

We want to identify key pulmonary lesions within the lungs of COVID-19 patients given a small set of labeled CT slice images. Creating pulmonary lesion masks is a complicated and time-consuming task that required radiology experts to perform it, making the availability of segmentation mask labels very limited. Previous approaches to solving this problem use individual models for each lesion, which often results in extreme over-fitting, especially for rarely present lesions. We propose to learn a single multi-task segmentation model to learn to segment all different lesions jointly instead. The use of multi-task learning also lets us take advantage of the information shared among the different lesion segmentation tasks for better performance and generalization. Our proposed “multi-task multidecoder segm. net” is inspired by the U-Net (Ronneberger, Fischer, and Brox 2015) architecture where the encoding part is shared among the different pulmonary lesion tasks with independent decoding heads. We refer to “multi-task segm. net” to a U-Net like multitask network architecture where both encoder and decoder parameters are shared among the different segmentation tasks. See Figure 5 describing the proposed network architecture on the Appendix section.

#### Multitask Segmentation Criterion

Both “multi-task multidecoder segm. net” and “multi-task segm. net” networks can be trained end-to-end using gradient-based optimization. The full criterion is described in Equation 4, where *α*_1_, *α*_2_, *α*_3_, and *α*_4_ are hyper-parameters. Since the task are very imbalance we use the ratio of the number of slices not showing the lesion over the number of slices showing that particular lesion as the corresponding alpha value. *L*_*GGO*_, *L*_*cl*_, *L*_*fl*_, and *L*_*it*_ can be any standard segmentation loss as Jaccard or binary cross-entropy (BCE). For our experiments we use weighted BCE with weights 0.3 for background and 0.7 for lesion prediction since often the lesions cover a very small region of the slices. Refer to the Appendix section for more implementation details.

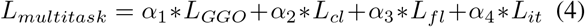

#### Lung Contour Segmentation

All segmentation slices have labels for lung contours and most standard semantic segmentation networks can segment the lung very accurately. For our pipeline, we used the U-Net architecture (Ronneberger, Fischer, and Brox 2015) to obtain a lung segmentation model with 94.09% mean IoU performance.

### 4.3 Multitask Segmentation Results

Table 5 shows the quantitative performance of our proposed approach compared to using individual models. Multitask models show better performance for all pulmonary lesions even when the number of trainable parameters was the same as the parameters of individual models. The performance improvement is even more noticeable in the low data regime as it is shown in Table 6 where models were trained using 50 percent of the training data ^1^. Figure 4 show qualitative results for the “multi-task multidecoder segm. net” on the test set. Model’s prediction closely align with the masks generated by expert radiologists.

**Table 5:** Lesion Segmentation Performance on CC-CCII Dataset

| Method | Ground-Glass |  | Consolidation |  | Fibrosis |  | Thickening |  | Num. of Params |
| --- | --- | --- | --- | --- | --- | --- | --- | --- | --- |
|  | mIoU (%) | GIoU (%) | mIoU (%) | GIoU (%) | mIoU (%) | GIoU (%) | mIoU (%) | GIoU (%) |  |
| Ground Glass Segm. Net. | 72.81 ± 0.07 | 55.94 ± 2.40 | -- | -- | -- | -- | -- | -- | 3.72 M |
| Consolidation Segm. Net. | -- | -- | 81.24 ± 1.80 | 83.14 ± 6.01 | -- | -- | -- | -- | 3.72 M |
| Fibrosis Segm. Net. | -- | -- | -- | -- | 89.88 ± 2.62 | 94.52 ± 7.28 | -- | -- | 3.72 M |
| Thickening Segm. Net. | -- | -- | -- | -- | -- | -- | 100.00 | 100.00 | 3.72 M |
| Multi-task Segm. Net. | 68.21 ± 3.50 | 53.01 ± 3.50 | 92.75 ± 5.92 | 90.79 ± 8.73 | 91.10 ± 2.80 | 95.34 ± 2.54 | 99.61 ± 0.32 | 100.00 | 3.72 M |
| Multi-task multidecoder Segm. Net. | **73.08 ± 2.59** | **74.225 ± 0.95** | 80.84 ± 0.98 | 84.22 ± 3.16 | 90.21 ± 2.15 | **95.52 ± 3.58** | **100.00** | 98.87 ± 1.96 | 7.78 M |

**Table 6:** Lesion Segmentation Performance 50 percent of data

| Method | Ground-Glass |  | Consolidation |  | Fibrosis |  | Thickening |  |
| --- | --- | --- | --- | --- | --- | --- | --- | --- |
|  | mIoU (%) | GIoU (%) | mIoU (%) | GIoU (%) | mIoU (%) | GIoU (%) | mIoU (%) | GIoU (%) |
| Ground Glass Segm. Net. | 67.31 | 51.16 | -- | -- | -- | -- | -- | -- |
| Consolidation Segm. Net. | -- | -- | 74.32 | 77.27 | -- | -- | -- | -- |
| Fibrosis Segm. Net. | -- | -- | -- | -- | 100.0 | 100.0 | -- | -- |
| Thickening Segm. Net. | -- | -- | -- | -- | -- | -- | 76.37 | 76.91 |
| Multi-task multidecoder Segm. Net. | **67.62** | **51.73** | **77.21** | **82.32** | **100.0** | **100.0** | **83.50** | **84.29** |

**Figure 4:**
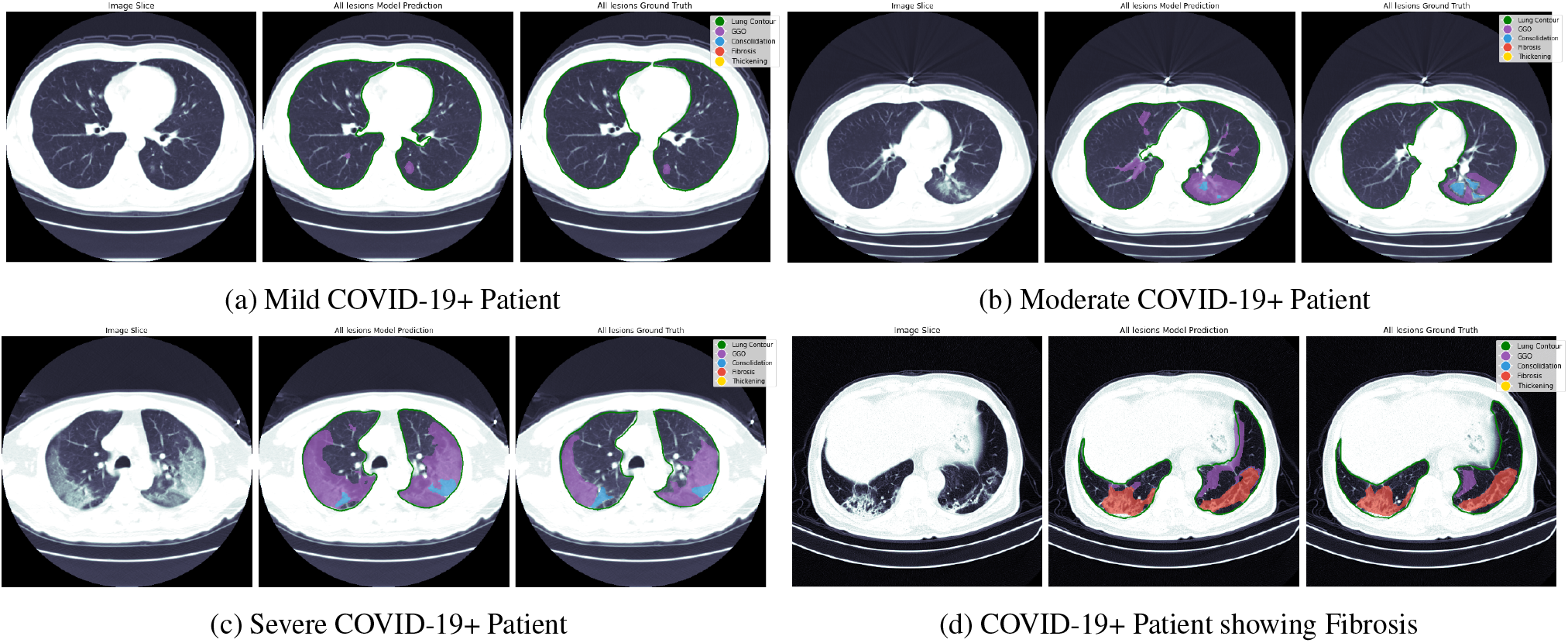
Qualitative results from our proposed lung segmentation and multitask segmentation network on COVID-19+ positive patients presenting different levels of disease severity. For every sub-figure the first image represents the input CT slice image, the second image represent our model prediction from all different lesions and the last image represent the segmentation ground truth obtained from expert radiologists. (a) Model predictions on a patient with mild novel COVID-19 pneumonia. This patient present signs of GGO (purple), (b) Model predictions on a patient with moderate novel COVID-19 pneumonia. This patient presents both GGO (purple) and consolidation (blue), (c) Model predictions on a patient with severe novel COVID-19 pneumonia, (d) Model predictions on a patient presenting severe pulmonary fibrosis (red) and GGO (purple).

**Figure 5:**
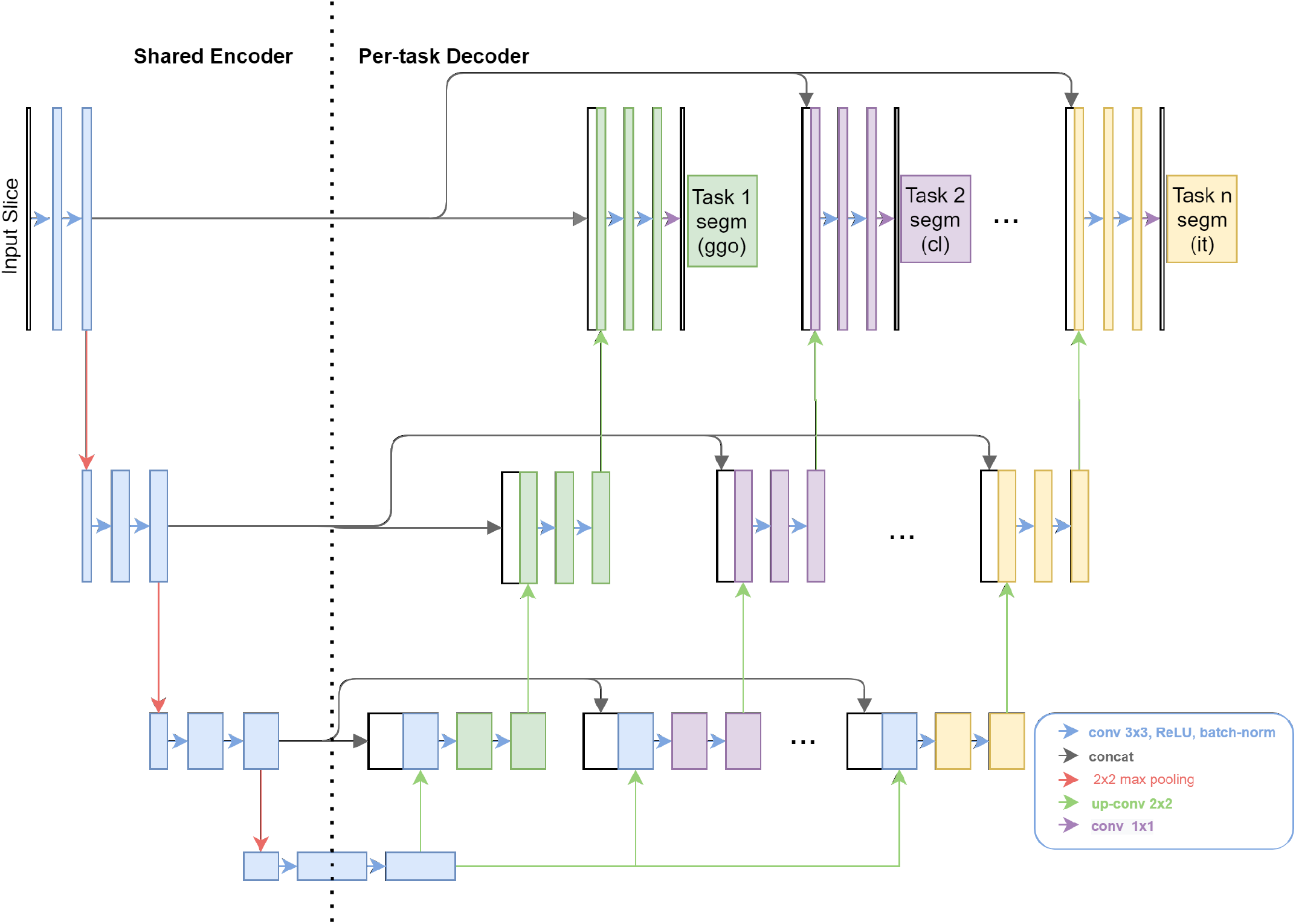
Diagram of our proposed multitask segmentation network (Best seen in color).

### 4.4 Lesion Anatomic Extent Estimation

A lesion anatomic extent is computed at the patient level as the percentage of the lung coverage by the lesion. The lung area is obtained using the lung segmentation model and the pulmonary lesion area is obtained from the multitask segmentation model predictions. For patients with multiple CT scans available, the final lesion score is obtained by averaging over the scan’s lesion scores. The anatomic extent of these pulmonary lesions will be used as features for the prognosis model presented in the following section.

## 5 Prognosis Model

In the CC-CCII dataset, a set of 61,810 CT slices from 436 COVID-19+ patients at hospital admission were labeled as severe and were not used for any of the experiments above. Patient demographic data (age, sex) were available for a subset of these patients as well. The data from 436 patients were combined with available demographics information to have a set of 105 patients with demographics and imaging feature data. We evaluated whether the pulmonary information obtained from these CT images using the feature extraction methods described above could be combined with demographic data to predict mortality. We compared the performance of models using demographic information alone, intensity map classification alone, segmentation alone, and combinations of classification and segmentation approaches combined with demographic information, in order to determine which model most accurately predicted mortality.As there were only 4 deaths recorded in the 105 patients, we applied sub-sampling on the majority class (survivors) to balance the dataset (40 survivors and 4 deaths). We performed prognosis experiments following the leave one participant out cross-validation (LOOCV), an extreme version of k-fold cross-validation where k is set to the number of examples in the dataset.

Extra randomized trees were trained to predict mortality using a combination of different feature sets (Table 7). The demographics alone (age and gender) achieved an AUC of 0.52 *±* 0.01. To explore the global feature set from the classification pipeline, we examined two separate cutoffs for features by PCA: the top 3 and the top 18 features, which explained 50% of the variance. The model using the top 18 PCA features performed poorly compared to using the top 3 features alone, indicative of having relatively few examples. Interestingly using local data from the segmentation pipeline alone achieved similar performance to using demographics data alone. However by including the segmentation features with the demographics, the AUC improved to 0.71 suggesting that imaging features encoded clinically relevant features for mortality prediction.

**Table 7:**
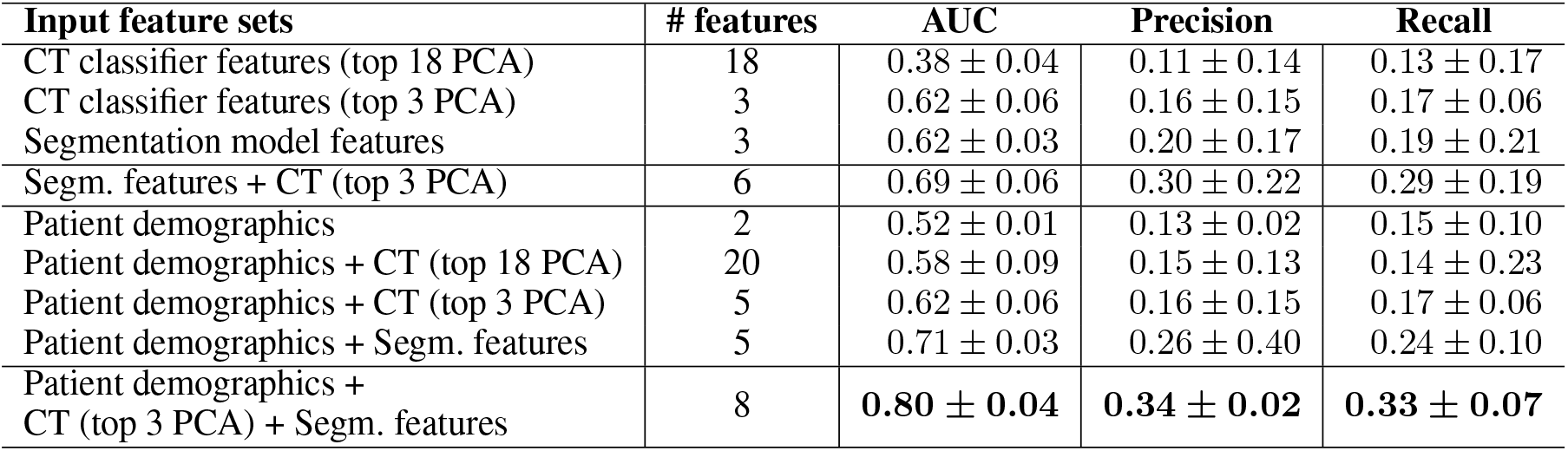
CC-CCII Prognosis Results with Leave One Out cross-validation

| Input feature sets | # features | AUC | Precision | Recall |
| --- | --- | --- | --- | --- |
| CT classifier features (top 18 PCA) | 18 | $0.38 \pm 0.04$ | $0.11 \pm 0.14$ | $0.13 \pm 0.17$ |
| CT classifier features (top 3 PCA) | 3 | $0.62 \pm 0.06$ | $0.16 \pm 0.15$ | $0.17 \pm 0.06$ |
| Segmentation model features | 3 | $0.62 \pm 0.03$ | $0.20 \pm 0.17$ | $0.19 \pm 0.21$ |
| Segm. features + CT (top 3 PCA) | 6 | $0.69 \pm 0.06$ | $0.30 \pm 0.22$ | $0.29 \pm 0.19$ |
| Patient demographics | 2 | $0.52 \pm 0.01$ | $0.13 \pm 0.02$ | $0.15 \pm 0.10$ |
| Patient demographics + CT (top 18 PCA) | 20 | $0.58 \pm 0.09$ | $0.15 \pm 0.13$ | $0.14 \pm 0.23$ |
| Patient demographics + CT (top 3 PCA) | 5 | $0.62 \pm 0.06$ | $0.16 \pm 0.15$ | $0.17 \pm 0.06$ |
| Patient demographics + Segm. features | 5 | $0.71 \pm 0.03$ | $0.26 \pm 0.40$ | $0.24 \pm 0.10$ |
| Patient demographics +<br>CT (top 3 PCA) + Segm. features | 8 | **$0.80 \pm 0.04$** | **$0.34 \pm 0.02$** | **$0.33 \pm 0.07$** |

The highest performing model used a combination of patient demographics, the top 3 PCA features from the classification model, and the segmentation features, for an AUC of 0.80 *±* 0.04. These results suggest that both the intensity mapping approach and the segmentation approach extract complementary clinical information from the CT volumes which, when considered together, are useful for predicting mortality. This is explainable as both the methods are addressing specific problems associated with the available training data. The intensity mapping allows for a global representation of features from the entire volume, while the segmentation model allows for the extraction of local but highly relevant features.

## 6 Conclusions

In this work we present two methods for automated extraction of clinically relevant features from chest CT images in the setting of limited data. We propose an intensity map projection method to create fixed size 2D representations of 3D CT volumes to facilitate COVID-19 diagnosis as well as a multitask segmentation network to extract highly relevant local features. These methods provide a way to integrate relevant information about pulmonary pathology (both global involvement and types of lesions) with other clinical data when creating prognosis models, and we demonstrate this by developing a model to predict mortality outcomes based on a combination of extracted CT data and patient demographic information. We show that when the CT features obtained using these methods are combined with patient demographics, the model is better at predicting mortality outcomes than when using demographic information alone.

## 7 Ethics Statement

The COVID-19 pandemic has created an urgent need for innovative approaches to diagnosis and clinical management, and the machine learning prognostic model proposed here could help allocate limited resources and may enable clinicians to provide earlier treatment interventions to prevent or mitigate severe disease. However, these potential benefits have not been validated in a clinical setting, and the proposed model should be evaluated in a real-world environment using data from diverse patient populations before clinical implementation. In addition, clinicians should have a clear understanding of how the model works before incorporating model predictions into their clinical decision-making. Patient consent is another ethical consideration that must be addressed with the use of prognostic models for triage or other clinical management.

## Data Availability

The data used for this work have been made publictly available by the China National Center for Bioinformation. http://ncov-ai.big.ac.cn/download?lang=en
The diagnosis ResNet model used pre-trained weights from Imagenet (http://www.image-net.org/)

http://www.image-net.org/

## A Multitask Segmentation Network Architecture

Figure 5 shows a detailed diagram of the proposed network architecture. It is a U-Net (Ronneberger, Fischer, and Brox 2015) like segmentation network with shared encoder and a decoder per class. There is flexibility in the depth of the network and the number of filters per layer. It is also possible to share parameters across the different tasks on the decoder as shown on the experiments in the the main document.

## B Segmentation Models Implementation Details

### Lung segmentation

To obtain the lung contour model we trained several U-Net models using resized to 512 *×* 512 input CT slices with different depths and number of input filters. We used the Adam optimizer (Kingma and Ba 2014) with a batch size of 24. All networks were trained from scratch with a learning rate of 0.001. We decay the learning rate by 10% on plateaus with patience of 5 epochs. The lung segmentation network was trained for 20 epochs. Weighted binary cross-entropy was used as the loss function. The best performing model on the validation set was picked as the final one.

### Multitask segmentation of pulmonary lesions

Both “multi-task multidecoder segm. net” and “multi-task segm. net” networks were trained end-to-end using Adam optimizer. The loss function used for training is defined in Equation 4. For *α*_1_, *α*_2_, *α*_3_, and *α*_4_ we use the ratio of the number of slices not showing the lesion over the number of slices showing that particular lesion as the corresponding alpha value. For *L*_*GGO*_, *L*_*cl*_, *L*_*fl*_, and *L*_*it*_ we used weighted binary cross-entropy with class weights 0.3 for background and 0.7. Both “multi-task multidecoder segm. net” and “multi-task segm. net” were trained from scratch with a learning rate of 0.001. We decay the learning rate by 10% on plateaus with patience of 5 epochs and train until convergence. A single training sample consist of a CT slice image along with a binary mask per task. We applied percentile standardization to each slice using the 2^*nd*^ and 98^*th*^ perecentiles. Slices where normalize to be zero-mean and show unit variance using mean and variance from the training split.

### Code

The PyTorch implementation of our proposed multitask segmentation network will be released on GitHub.

## Footnotes

1 Number of available test slices for fibrosis and thickening is very small and might not reflect accurately model’s performance

## Notes

### Competing Interest Statement

The authors have declared no competing interest.

### Funding Statement

WCL was supported by unrestricted research funds to WCL from the Department of Medicine, University of Washington.

### Author Declarations

The data used on this paper is publicly available and anonimyzed.

